# Risk factors for patients with social determinants of health not to follow up with community-based organizations to which they have been referred

**DOI:** 10.64898/2026.02.28.26347084

**Authors:** Rida Nasir, Aleeza Nasir, Diamia Foster, Daisy Puca, Kevin Charles, Mark Richman

**Affiliations:** Hofstra University; Johns Hopkins; Binghamton University; Northwell Health Long Island Jewish Medical Center

## Abstract

This study explores the influence of social determinants of health (SDOH) on follow-up behavior among patients referred to community-based organizations (CBOs) in the Emergency Department (ED) of Long Island Jewish (LIJ) Medical Center. A retrospective analysis was conducted on data collected from 342 patients who were screened for SDOH between February and July 2023. Descriptive statistics and Chi-squared tests were used to identify potential associations between demographic and social factors (race, language, age, gender, employment status, and insurance status) and follow-up rates. The results revealed several trends: non-White patients (73.2%) and non-English speakers (81.8%) followed up more frequently than their counterparts, as did older adults (80.0%) and insured patients (77.8%). However, none of the variables reached statistical significance (all p-values > 0.05). The findings suggest that while demographic and social factors may influence follow-up behavior, the lack of statistical significance could be attributed to the limited sample size. These trends align with previous literature on SDOH and follow-up behavior, highlighting the need for further research with larger, more representative samples. Addressing the complex interplay of SDOH, including factors such as language, insurance, and cultural differences, is crucial for improving follow-up rates and ensuring better health outcomes for underserved populations. Future research should focus on refining referral systems, exploring additional socioeconomic factors, and conducting longitudinal studies to develop more effective strategies for integrating SDOH interventions in healthcare systems.

## Introduction

Social determinants of health (SDOH) can significantly influence health outcomes, with limited access to necessary resources often contributing to higher hospitalization rates and increased healthcare costs. Lack of access to medical care can exacerbate health disparities and hinder individuals’ abilities to manage chronic conditions, maintain preventive care, and improve overall well-being (Krause *et al*., 2021). The CDC maintains that SDOH factors have a greater impact on health outcomes than genetic or environmental factors alone (CDC, 2024).

Chronic diseases are closely linked to SDOH, as unmet social needs often exacerbate disease progression and reduce patients’ abilities to manage their conditions effectively. A study investigating the relationship between social determinants of health and functional capacity in adult primary care patients with multiple chronic conditions found that patients experiencing any form of SDOH burden, such as financial, food, or housing insecurity, demonstrated significantly lower functional capacity, even after adjusting for social and demographic factors (Bonnell *et al*., 2021). Social determinants of health (SDOH) play a critical role in shaping mental health outcomes, with identity factors such as race, gender, and sexual orientation influencing the risk of mental health disorders like major depressive disorder (MDD). A study examining the role of SDOH in mental health found that food insecurity and loneliness significantly increased the likelihood of MDD, while identity characteristics, such as gender and LGBTQIA2+ status, contributed to disparities in depression risk, with women and LGBTQIA2+ individuals having notably higher odds (Kammer-Kerwick *et al*., 2024). Addressing social determinants of health (SDOH) through targeted interventions in areas such as education, early childhood development, urban planning, housing, income support, and employment has been shown to reduce health disparities (Thornton *et al*., 2016). Additionally, a recent study examining SDOH effects on hospitalization rates found that adults with multiple SDOH burdens, such as food insecurity, social isolation, and low educational attainment had a significantly higher risk of hospitalization (Wray *et al*., 2022). Another study found that Medicaid beneficiaries lacking adequate social or community support, those who frequently felt isolated, incurred an average of $2,707 higher annual healthcare costs (Mohan & Gaskin, 2024). The impact of SDOH on health outcomes and healthcare costs suggests a need for prioritizing interventions that tackle these factors in order to foster a more equitable and efficient healthcare system (Crear-Perry *et al*., 2021).

### Key Domains of SDOH

Social determinants of health (SDOH) encompass several key domains, including education, housing, employment, food security, healthcare access, social support, and neighborhood conditions (American Academy of Family Physicians, 2019). These factors shape individuals’ health by influencing their access to essential resources and opportunities. Lower levels of education have been shown to correlate with worse health outcomes, including higher rates of chronic diseases and reduced life expectancy (Zajacova & Lawrence., 2024). A study found that individuals with lower education levels report poorer health outcomes, contributing to increased incidences of conditions such as hypertension, diabetes, and obesity (Kubota *et al*., 2017). Additionally, health literacy, which is directly related to education, affects individuals’ ability to manage chronic diseases, such as diabetes (Seiglie *et al*., 2020).

Employment also plays a crucial role in health by providing financial stability and access to healthcare (Posel *et al*., 2021). Unemployment has been linked to higher levels of stress, poor mental health, and a greater risk of chronic conditions (Picchio & Ubaldi, 2024). Research has shown that unemployed individuals are more likely to report poor mental health and are at an increased risk of conditions such as heart disease (Machicado *et al*., 2017). Moreover, job-related stress is recognized as a risk factor for cardiovascular diseases (Wang *et al*., 2021). Housing conditions significantly influence health outcomes, as safe, stable housing promotes better health and access to healthcare resources (Rolfe *et al*., 2020). Poor housing conditions such as overcrowding, inadequate heating, and exposure to environmental hazards can lead to increased risks of respiratory diseases and mental health problems (Marí-Dell’Olmo *et al*., 2017). Inadequate housing quality is also associated with a higher incidence of asthma and mental health disorders such as anxiety and depression (Baker *et al*., 2016).

Food insecurity functions as a critical determinant of health by influencing disease prevention, management, and overall well-being (Hanmer *et al*., 2021). Individuals experiencing food insecurity are more likely to have poor nutrition, contributing to chronic conditions like diabetes, obesity, and cardiovascular disease (Thomas *et al*., 2021). Furthermore, food insecurity exacerbates existing health disparities, particularly among low-income populations (Wolfson *et al*., 2020). Social support, including emotional support from family, friends, and community, plays a significant role in promoting mental and physical health (Kelly *et al*., 2017). Studies have shown that individuals with strong social networks have better health outcomes, indicating a positive association between higher levels of social support and lower levels of inflammation (Lee *et al*., 2019). Social support has been found to counteract the negative effects of stress, contributing to better health outcomes in both chronic disease management and prevention (Liu *et al*., 2021).

Finally, the quality of the neighborhood where individuals reside significantly impacts their Quality-Adjusted Life (QAL) status. For instance, research shows correlation to exist between higher neighborhood socioeconomic status with post-stroke recovery status (Twardzik *et al*., 2019). Conversely, another study found that neighborhoods with lower socioeconomic status showed the greatest yield in mental health benefits from increased greenspace (Brown *et al*., 2018). Research indicated a strong influence of neighborhood conditions on QAL status, suggesting that improvements be targeted to underserved and low-income areas to improve mental and physical health. Collectively, these domains created a framework in which disparities in access to resources can lead to inequitable health outcomes (Kneipp *et al*., 2018). Addressing these factors through policy changes and interventions can significantly reduce health disparities and enhance the overall well-being of vulnerable populations (Brown *et al.,* 2019).

### Various Determinants of Health and Healthcare Utilization Patterns

Social determinants of health significantly influence healthcare access and utilization, with extensive research demonstrating that underserved populations are at higher risk for chronic diseases, including diabetes (Hill-Briggs *et al*., 2020). Among these determinants, housing stability plays a particularly critical role, directly shaping healthcare utilization patterns in older adults. Factors such as housing ownership and housing-related challenges influence both the likelihood and intensity of home care service use in this population (Mah *et al*., 2021).

Furthermore, a study examining the impact of housing instability on the healthcare patterns of caregiver and child health among low-income renter families found that housing instability-such as being behind on rent, frequent moves, and homelessness-was associated with negative health outcomes, indicating an increased need for healthcare utilization services (Sandel *et al*., 2018).

Food insecurity, another critical social determinant of health, also significantly influences healthcare utilization patterns. Research has shown that limited access to adequate nutrition not only exacerbates chronic health conditions but also increases the need for healthcare services, with individuals reporting poorer overall health and increased utilization of emergency department services (Berkowitz *et al*., 2018). Another study corroborated these findings through an investigation of food insecurity among pediatric patients (Thomas *et al*., 2019). Moreover, access to healthcare, particularly through health insurance as a key determinant, can influence how individuals engage with healthcare services. For example, a study analyzing healthcare expenditures and usage revealed that the ACA’s young adult expansion led to an increase in office-based visits, a decline in emergency department visits, and a reduction in overall healthcare costs (Deb & Norton, 2018). Another study indicated that physical multimorbidity was linked to higher rates of outpatient visits, hospitalizations, and substantial financial burden from healthcare costs, suggesting the need for improved health insurance coverage (Zhao *et al*., 2020).

Educational attainment, another significant determinant of health, can influence healthcare utilization, as knowledge of one’s condition and the importance of preventive care is crucial for effective health management. A lack of understanding may deter individuals from seeking care, particularly if they do not fully comprehend the severity of their illness or the necessity of preventive measures. A study investigating the impact of education on the utilization of health services in a rural population in Ghana found educational level positively correlated with healthcare utilization (Agyemang & Asibey, 2018). Researchers corroborated these results when examining factors influencing health service utilization for common mental disorders, finding that higher educational levels were linked to healthcare use (Roberts *et al*., 2018).

Race, as a social determinant of health, can also impact healthcare utilization, with minority and underserved communities experiencing the greatest burden. A systematic review investigating disparities in healthcare utilization among adults with traumatic brain injuries found that racial and ethnic minorities were less likely to utilize both in-hospital and post-hospital healthcare services, while other studies did not find a significant link between race/ ethnicity and healthcare utilization among these patients (Gao *et al*., 2018). Within the emergency department setting, a study found that Black and Hispanic patients had higher utilization rates than their white counterparts (Parast *et al*., 2022). The complex relationship between social determinants and healthcare utilization highlights the varying degrees to which they influence one another.

### Economic Implications of Addressing Social Determinants of Health (SDOH) in Healthcare

Moreover, failure to address fundamental SDOH needs can lead to increased healthcare spending from both consumer and payer perspectives. In 2022 alone, national health expenditure data reported a budget of $4.5 trillion. Projections estimate that the share of national health expenditure (NHE) in GDP will reach 19.7% by 2032, as NHE growth is expected to consistently outpace national GDP growth (Centers for Medicare & Medicaid Services, 2024). Addressing SDOH needs could lead to lower healthcare utilization rates, which in turn may help offset the significant rise in spending.

Research shows that housing instability contributes to higher healthcare encounter rates, a trend that is exacerbated in neighborhoods with lower SES (Hatef, 2021). Among commercially insured individuals, this trend is also evident, with higher disease prevalence, increased emergency room utilization, and higher healthcare spending, with individuals incurring approximately $1,200 per person per year (Falconi *et al*., 2023). Unmet social needs affect individuals across all groups, with research showing that consumers, regardless of their level of unmet social needs, report similar patterns. The findings revealed that those with higher healthcare utilization rates were also more likely to report unmet SDOH needs (Coe *et al*., 2020). This particularly places additional strain on emergency care, which is already experiencing a shortage of physicians, as the growing reliance on emergency services is likely to intensify the existing burden.

A study investigating the association between SDOH documentation and Emergency Department admission found that patients with documented needs were more likely to be admitted to the hospital through the ED (McCormack & Brown, 2021). This not only exacerbates the strain on already overburdened ED resources but also drives up healthcare expenditures. Patients with documented SDOH needs often face barriers to accessing primary or preventive care, resulting in increased reliance on the ED (Ryus *et al*., 2023). This increased utilization subsequently leads to higher treatment costs, thereby contributing to the financial burden on the healthcare system.

Strains on the healthcare system are intensified by inadequate screening for SDOH. Studies reveal operational inefficiencies within current EHR systems and highlight the underutilization of IT tools for screening (Hatef *et al*., 2024). Better documentation of SDOH needs can allow for a reduction in healthcare cost. Implementing better documentation of SDOH needs within clinical settings is crucial for identifying and addressing unmet social needs. This can help to mitigate some of the financial burden of rising healthcare costs by reducing expenses associated with avoidable healthcare utilization. By integrating comprehensive SDOH data into clinical workflows, healthcare systems can also allocate resources more efficiently and prevent resource misallocation (Kirksey *et al*., 2024).

This lack of effective screening worsens the burden of unmet SDOH needs, further contributing to higher costs and the misallocation of resources, ultimately reflecting systemic inefficiencies (Sezgin *et al*., 2024). Predictive analytics combined with AI-driven SDOH programs enable earlier identification of high-risk patients who are more vulnerable to specific needs. These programs improve resource allocation by tailoring interventions to address individual concerns, thereby reducing costs by minimizing unnecessary visits and avoiding unnecessary procedures (Nassif & Khalili, 2024).

From the perspective of healthcare institutions, AI-driven SDOH programs contribute to optimizing bed utilization by effectively addressing less emergent patient needs. These technologies facilitate more timely interventions, thereby reducing the demand for hospital resources and improving overall operational efficiency. Addressing SDOH needs is an essential step in mitigating disparities and inequities within healthcare, as well as in reducing expenditures within an already strained budget. It also underscores the ongoing healthcare crisis, characterized by inefficiencies in spending, as significant needs remain unmet.

In light of the growing trend of consumerism, the findings indicate that, despite substantial investment in healthcare from a payer perspective, merely increasing the healthcare budget without addressing underlying factors, such as social determinants of health, does not result in a more effective utilization of healthcare resources. In addition to addressing the direct healthcare costs and utilization rates associated with SDOH, the broader economic implications of such interventions must also be considered. Social Return on Investment (SROI) offers a framework to measure these broader impacts, providing insights into the value created for society at large, including reduced social disparities and long-term market benefits. However, research on the link between SDOH screening and SROI remains limited, with studies indicating mixed impacts on metrics such as utilization and costs (Yan *et al*., 2022) & (Chuang *et al*., 2023).

### Integrating Community-Based Organizations and Referral Platforms to Address SDOH

To address unmet needs effectively, healthcare infrastructure must evolve to incorporate community-based organizations (CBOs) as central players in addressing social determinants of health (SDOH). Community-Based Organizations (CBOs) are nonprofit, grassroots, or local organizations that operate within communities to address various social, economic, and health-related challenges. These organizations are uniquely positioned to understand and address the specific needs of their communities (Hussain, 2024). CBOs often provide services such as housing assistance, food security programs, education and job training initiatives, and access to healthcare resources (Engelman *et al*., 2022).

In the context of healthcare, CBOs play a vital role in mitigating SDOH by bridging the gap between clinical settings and social support networks. By addressing barriers such as housing instability, food insecurity, and access to transportation, CBOs help reduce healthcare utilization and improve outcomes for underserved populations (Kranz *et al*., 2020). The capacity of CBOs to deliver targeted interventions and community-specific resources positions them as essential contributors to fostering a more equitable healthcare system (Meigs *et al*., 2024). Referral platforms are essential in improving access to these community resources and enhancing the effectiveness of CBOs in addressing SDOH (Crumley *et al*., 2023). Settings like emergency departments can disproportionately serve low-income and patients, making them a vital point for intervention (Samuels-Kalow *et al*., 2021).

A study exploring the integration of a social needs screening tool and referral process through an academic-community partnership demonstrated the utility of CBO resources, such as pharmacy programs and utility assistance, facilitated by existing tools like the United Way 2-1-1 system. Results found that the integration of CBOs facilitated the identification of SDOH-vulnerable patients, with nearly half of those requesting referrals successfully connecting to community resources (Wallace *et al*., 2020). Another study on the implementation of an EHR-based screening and referral system in primary care found that automating referrals to community resources streamlined the process for healthcare providers, facilitating better support for patients’ social determinants of health (de la Vega *et al*., 2019).

While the integration of CBOs and referral platforms have shown promise in addressing social determinants of health, several barriers can hinder patients from successfully following through with referrals to these essential resources. For instance, a study investigating barriers to enrollment in community-based physical activity programs finds that cost, time constraints, and poor referral processes hinder participation, while motivation and social support encourage it (Schumacher *et al*., 2024). Another study exploring factors influencing follow-up in a community-based rehabilitation program finds that gaps in post-program services, unclear referral processes, poor communication among care settings, and program structure constraints hinder follow-up, leaving participants feeling unsupported (Maximos *et al*., 2024).

While barriers and facilitators influence follow-up rates to community-based resources, the limited adoption of referral platforms also highlight additional systemic challenges that hinder the integration of technology into these processes. For instance, research on factors influencing use of a community resource referral platform reveals low adoption due to perceived lack of utility and technological challenges (Fichtenberg *et al*., 2024). Another study investigating factors affecting the adoption of web-based social service referral technology in the nonprofit sector supports these findings, identifying resource constraints, technology compatibility, and perceived usefulness and efficiency as key factors contributing to the low adoption of such programs (Hutton, 2022).

### Purpose

Addressing the social determinants of health (SDoH) has become an essential focus of healthcare institutions aiming to reduce disparities and improve patient outcomes. Northwell Health at Long Island Jewish (LIJ) Medical Center previously undertook a study to address this challenge by implementing an Emergency Department Discharge Center (EDDC). Established in July 2020, the EDDC aimed to enhance follow-up care and increase referrals to community-based organizations (CBOs) for patients screened for SDoH during emergency department (ED) visits (Razack *et al*., 2024). This initiative involved screening patients using a detailed form, entering data into a social services portal, and providing information about local CBOs via Unite Us® (formerly NowPow®). Despite these efforts, their internal review highlighted significant challenges, including a follow-up rate of only 4.5% among referred patients (Razack *et al*., 2024).

While the EDDC demonstrated a commitment to addressing SDoH, key challenges persisted. During its pilot year, only 1% of ED discharges were screened for SDoH, though efforts have since increased to a projected 6.7% of discharges (Razack *et al*., 2024). These findings emphasize the need to examine patient-level risk factors, such as housing status, zip code, race, language, age, gender, employment status, and insurance coverage, that may hinder follow-up with CBOs. Identifying these factors can guide the development of tailored strategies to improve patient engagement and outcomes.

Building upon this foundation, this study investigates the specific risk factors associated with low follow-up rates among patients identified with one or more SDoH burdens. By leveraging the data made available through LIJ Medical Center’s EDDC, this research seeks to provide recommendations for ED discharge staff to address these vulnerabilities effectively, thereby bridging the gap between patients and essential community resources.

## Methodology

This study was conducted as a retrospective analysis of data collected from 342 patients who were offered screening for social determinants of health (SDOH) in the Emergency Department (ED) of Long Island Jewish (LIJ) Medical Center between February and July 2023. LIJ is a 583-bed hospital serving a racially and socioeconomically diverse population, including a significant proportion of uninsured or underinsured patients. The adult ED sees approximately 100,000 patients annually and discharges around 75,000 patients (Razack et al., 2024). Northwell’s service area, which includes LIJ, encompasses some of the most racially, ethnically, and linguistically diverse communities in the United States, spanning urban, suburban, and rural settings. A large immigrant population, primarily Spanish- and Bengali-speaking, is also served (Northwell Health, n.d.).

### Data Collection

The data was sourced from a database that included information on patients’ SDOH screening and follow-up outcomes. The screening was conducted using a paper-based survey, and patients were referred to community-based organizations (CBOs) through Unite Us® / NowPow, an electronic referral platform. The database captured patient identifiers and demographics, including race, language, age, gender, employment status, and insurance status. Additionally, the database recorded whether patients opted to complete the survey, identified one or more SDOH needs, and followed up within two weeks with a referred CBO. Housing status, employment status, reported mental illness, and gender were recorded as binary variables. Insurance status was categorized as Medicare, Medicaid, private, or uninsured (none/self-pay). In cases where patients had both Medicare and Medicaid, Medicare was prioritized in the dataset. Missing data were excluded from the analysis.

This study was deemed exempt by the Northwell Health Human Subject Protection Program - Institutional Review Board as a quality improvement project. All data were anonymized to protect patient confidentiality, and the analysis was conducted in compliance with ethical standards.

### Data Analysis

To identify similarities and differences between patients who followed up with a community-based organization (CBO) and those who did not, descriptive statistics were employed. Percentages for each variable were calculated across both groups. A Chi-squared test with (N-1) degrees of freedom was used to determine significant associations between follow-up status and demographic/social factors. Statistical significance was set a priori at p < 0.05. The variables analyzed included race, language, age, gender, employment status, and insurance status. The follow-up rates and p-values for these variables are listed in Table 1.

**Table 1:** Follow-up rates by demographic and social factors among patients referred to community-based organizations (CBOs).

| **Demographic/Social Factor** | **Group** | **Follow-up Rate** | **p-value** |
| --- | --- | --- | --- |
| Race | White | 55.6% (5/9) | 0.285 |
|  | Non-White | 73.2% (41/56) |  |
| Language | English | 68.5% (37/54) | 0.3805 |
|  | Non-English | 81.8% (9/11) |  |
| Age | Over 64 | 80.0% (8/10) | 0.4891 |
|  | 64 and Younger | 69.1% (38/55) |  |
| Gender | Male | 64.5% (20/31) | 0.2917 |
|  | Female | 76.5% (26/34) |  |
| Employment Status | Employed | 71.4% (10/14) | 0.9539 |
|  | Unemployed | 70.6% (36/51) |  |
| Insurance Status | Insured | 77.8% (21/27) | 0.2982 |
|  | Uninsured | 65.8% (25/38) |  |

By examining these variables, the study aimed to identify patterns or factors associated with follow-up behaviors, contributing to a deeper understanding of how SDOH influences patient outcomes and the utilization of CBO resources.

## Results

Out of the 342 patients screened for social determinants of health (SDOH) and referred to community-based organizations (CBOs), follow-up rates revealed notable trends, though none of the variables reached statistical significance (all p-values > 0.05). Non-White patients followed up with CBOs more often (73.2%) compared to White patients (55.6%). Regarding language, non-English speakers had higher follow-up rates (81.8%) compared to English speakers (68.5%). Age-related differences showed that patients over 64 years old had higher follow-up rates (80.0%) compared to those aged 64 or younger (69.1%). Gender disparities were observed as well, with female patients following up more frequently (76.5%) than male patients (64.5%). Differences in employment status and insurance coverage were comparatively smaller. Employed patients followed up at a rate of 71.4%, similar to non-employed patients (70.6%). Likewise, insured patients had a higher follow-up rate (77.8%) than uninsured patients (65.8%). These trends suggest potential disparities in follow-up behavior influenced by demographic and social factors. However, further research with a larger sample size is needed to confirm these findings and assess their significance.

## Discussion

The analysis of follow-up rates among patients referred to community-based organizations (CBOs) revealed several trends. Non-White patients exhibited higher follow-up rates (73.2%) compared to White patients (55.6%), and non-English speakers also showed higher follow-up rates (81.8%) compared to English speakers (68.5%). Furthermore, older adults (80.0%) and insured patients (77.8%) demonstrated greater follow-up rates than their respective counterparts. Despite these trends, none of the demographic or social factors analyzed, including race, language, age, gender, employment status, or insurance status, were statistically significant (p-values > 0.05). This suggests that while certain patterns were observed, there was no strong statistical evidence to support the direct influence of these factors on follow-up behavior in this sample.

When comparing the findings of this study to existing literature, the observed trends are partially consistent with prior research on social determinants of health (SDOH) and follow-up behavior. Previous studies have consistently identified race, language, and insurance status as significant factors influencing follow-up behavior, although the degree of their impact can vary. Research has frequently indicated that non-White populations and older adults tend to engage more with community-based resources (Uwemedimo & May, 2018) & (Gibbons, 2022), a pattern that aligns with the findings of this study. For instance, while this study observed a higher follow-up rate among non-White patients, the absence of statistical significance may be due to the sample size or the presence of other unmeasured factors influencing follow-up behavior. Similarly, although language barriers were often cited as a challenge to follow-up, non-English speakers in this study exhibited higher-than-expected follow-up rates (Pang *et al*., 2023).

The lack of statistical significance may be related to the limited sample size, which could have impacted the ability to detect meaningful differences in follow-up rates. Larger sample sizes, with more complete data, would likely provide greater statistical power to identify associations between SDOH and follow-up behavior. Additionally, follow-up behavior is influenced by a complex array of factors, and smaller sample sizes may not capture the full range of variables that contribute to these outcomes.

Despite the lack of statistical significance, the observed trends highlight the complexity of social determinants of health and their interaction with individual patient circumstances. Factors such as language barriers, cultural differences, and varying levels of access to resources may influence follow-up behavior, but these effects can be confounded in smaller samples. Furthermore, systemic challenges in healthcare such as inconsistent screening for social determinants or difficulties in coordinating referrals may also contribute to the findings. These challenges emphasize the need for more targeted interventions to improve follow-up rates, particularly among underserved populations. Understanding the barriers and facilitators that impact follow-up, informed by larger and more representative samples, is essential for improving the effectiveness of community-based referrals and enhancing patient outcomes.

## Conclusion

The findings of this study underscore the importance of understanding the role social determinants of health (SDOH) play in influencing follow-up behavior among patients referred to community-based organizations (CBOs). Several trends emerged, such as higher follow-up rates among non-White patients, non-English speakers, older adults, and insured patients. These observations highlight potential areas for targeted intervention and suggest that factors such as language barriers, cultural differences, and insurance status may influence follow-up behavior.

Considering the public health implications of this study, addressing social determinants of health (SDOH) within healthcare systems is essential. This can be achieved through optimizing referral systems, enhancing resource accessibility, and developing tailored interventions specifically aimed at underserved populations. The limitations of this study suggest the need for larger studies to better understand the impact of SDOH on follow-up behavior and improve the effectiveness of related interventions.

Future research should explore additional variables that could influence follow-up behavior, such as mental health, transportation access, and other socioeconomic factors. Longitudinal, prospective studies could also provide deeper insights into how these factors interact over time, leading to more effective strategies for improving patient outcomes. Additionally, future studies should continue to focus on refining referral systems and integrating SDOH interventions into healthcare to ensure that all patients, particularly those from underserved communities, have access to the resources they need for disease prevention and health promotion.

## Data Availability

N/A

## Notes

### Competing Interest Statement

The authors have declared no competing interest.

